# Towards course of disease based epidemiological modelling: motivation and computational optimization

**DOI:** 10.1101/2023.05.24.23290318

**Authors:** Yu-Heng Wu, Torbjörn E. M. Nordling

**Affiliations:** Department of Mechanical Engineering, National Cheng Kung University, Tainan, Taiwan

**Keywords:** COVID-19, individual data, synthetic dataset, epidemiological model, firefly optimization

## Abstract

The ongoing COVID-19 pandemic has demonstrated the shortcoming of epidemiological modelling for guiding policy decisions. Due to the lack of public data on infection spread in contact networks and individual courses of disease, current forecasting models rely heavily on unreliable population statistics and *ad hoc* parameters, resulting in forecasts with high uncertainty. To tackle the problem of insufficient public individual data, we develop an agent-based model to generate a synthetic Taiwanese COVID-19 dataset. We collected COVID-19 data from Taiwanese public databases for the period when the original SARS-CoV-2 virus was most prevalent (Jan.-Oct., 2020) and fit our model to it. We used the Firefly algorithm to optimize the 194 epidemiological parameters and validated the synthetic dataset by comparing it to Taiwanese public data. Here we study the difference between population statistics and individual course of disease data, and computational optimization of our code to reduce run time. The discrepancy between serum prevalence and reported cases, as well as excess deaths and reported deaths, show that population statistics are unreliable. Monte Carlo simulations further exemplify the discrepancy between actual and reported infections. By using Python CProfiler and Snakeviz packages, we iteratively optimize our algorithm and has so far decreased the computation time of the core code from 0.11s to 0.07s. The large computation time implies that we need to further optimize the algorithm.

## I. Introduction

The COVID-19 epidemic outbreak caused by Severe Acute Respiratory Syndrome Coronavirus 2 (SARS-CoV-2) has spread worldwide since December 2019. The decision-making process was often based on mathematical modelling for forecasting and/or intervention comparison purposes. The UK’s first lockdown was based on Imperial College’s forecast [1]. Hellewell *et al*. has investigated the effect of quick isolation on controlling the pandemic [2]. They concluded that highly effective contact tracing and case isolation is enough to control a new COVID-19 outbreak. The Institute for Health Metrics and Evaluation (IHME) used the deterministic Susceptible-Exposed-Infected-Recovered (SEIR) model to provide forecasts under a variety of scenarios, such as mask use, vaccination, and antivirals use [3]. A trustworthy forecasting model is crucial to guide the disease prevention policy. However, current epidemiological forecasting models are based on population data, which require precise population measurements such as the accurate number of infected or deaths. Individual-level data is more informative from a modelling perspective but almost completely lacking in any public database, limiting the development of epidemiological models.

In general, COVID-19 models can be divided into (i) population-based models and (ii) individual-based models. On the population-based side, the model is either only based on population data or the concept of virus spread at the population level. Widely used population data are the daily number of confirmed cases and recovered. At the beginning of the pandemic, the Susceptible-Infected-Recovered (SIR) model SIKJ*α* was implemented, forecasting the temporally varying infection, death, and hospitalization rates [4]. Some other teams developed deep learning models to extract the hidden pattern in the population data. DeepCOVID created a small-sized deep neural network (DNN) due to the small dataset that included CDC data and syndromic surveillance data [5]. Microsoft’s deep learning model, DeepStia (HierST), considered not only the population cases but also spatial information [6]. They trained the model containing two graph neural networks (GNN) to explain the interaction between country, states, and county in the US. In the diffusion convolutional recurrent neural network—DeepGLEAM [7]—the encoder reads as input a 7 *×* 50 *×* 4 tensor that consists of the daily residuals between the observed death number and GLEAM forecast. The decoder produced forecasts for each state for the following 4 weeks.

The individual-based model (agent-based model) is based on the disease-spreading structure that incorporates the disease spread at the individual level (agent), *i*.*e*. how the virus spread from one person to another. Imperial College of London developed an agent-based stochastic model (CovidSim) and simulated the pandemic for the UK and USA [1]. The model can provide forecasting of cases, deaths, and hospitalisations under different non-pharmaceutical interventions such as suppression and mitigation. Institute for Disease Modeling (IDM) developed the agent-based model, COVID-19 Agent-based Simulator (Covasim), simulating each agent’s demographic data, disease progression, and contact network in different social layers [8].

Despite this, all models performed poorly with high uncertainty, and none of them was trained on individual data. We focus on developing a forecasting model based on individual-level course of disease datasets. We first develop an algorithm that generates an individual-level synthetic dataset. The main idea of the branching process of our algorithm is shown in Figure 1 and the detailed flow chart is shown in Figure 2.

**Fig. 1:**
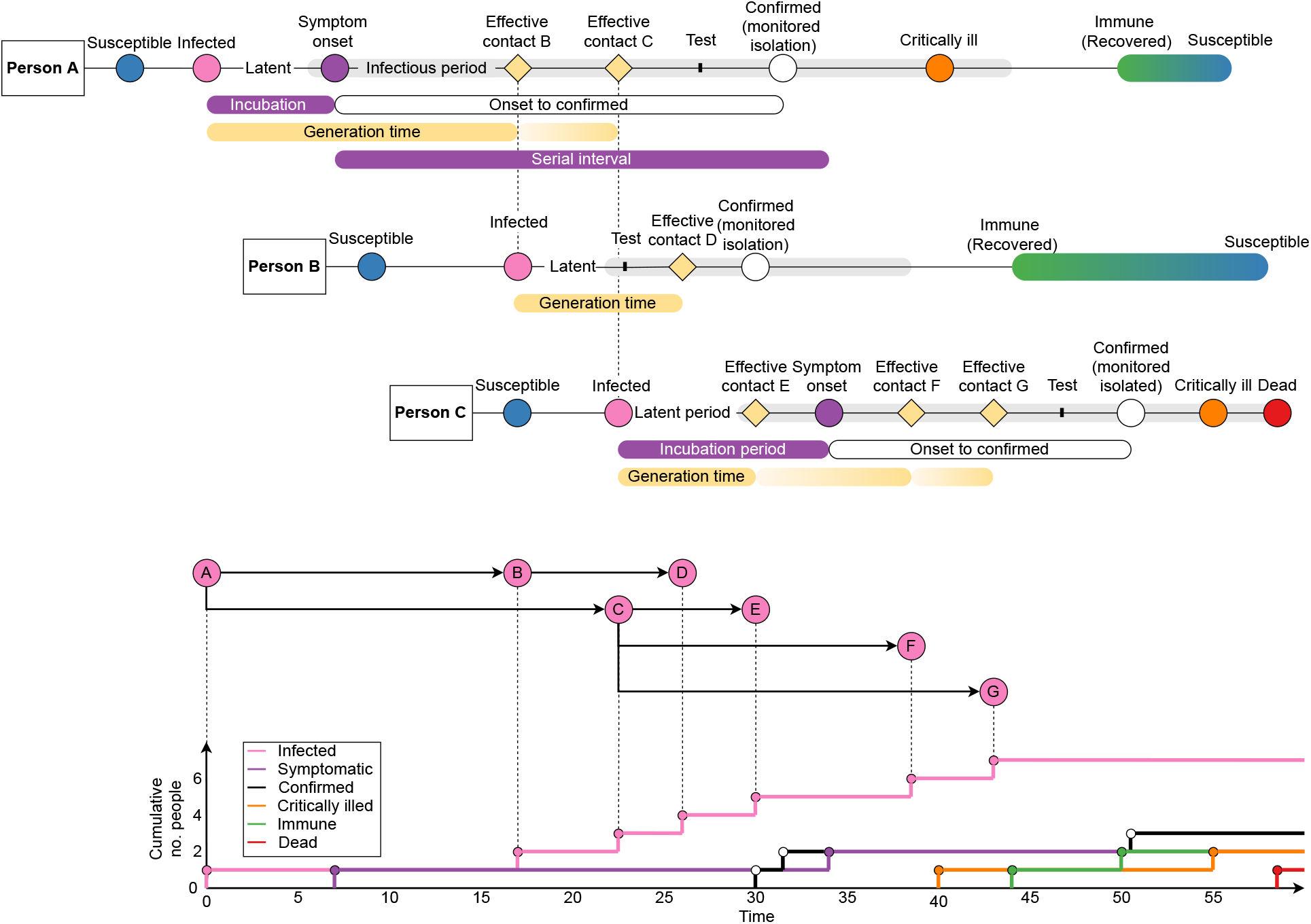
Example of the simulated branching process model and the mapping to the daily cumulative infected cases. Case B and C are the effective contacts of case A, case D get infected by case B, and case E to G are infected by case C. The bottom plot shows that the resulting chains can be mapped into cumulative cases per day, which follow an exponential growth trend. Reproduced from a manuscript under CC BY license [9].

**Fig. 2:**
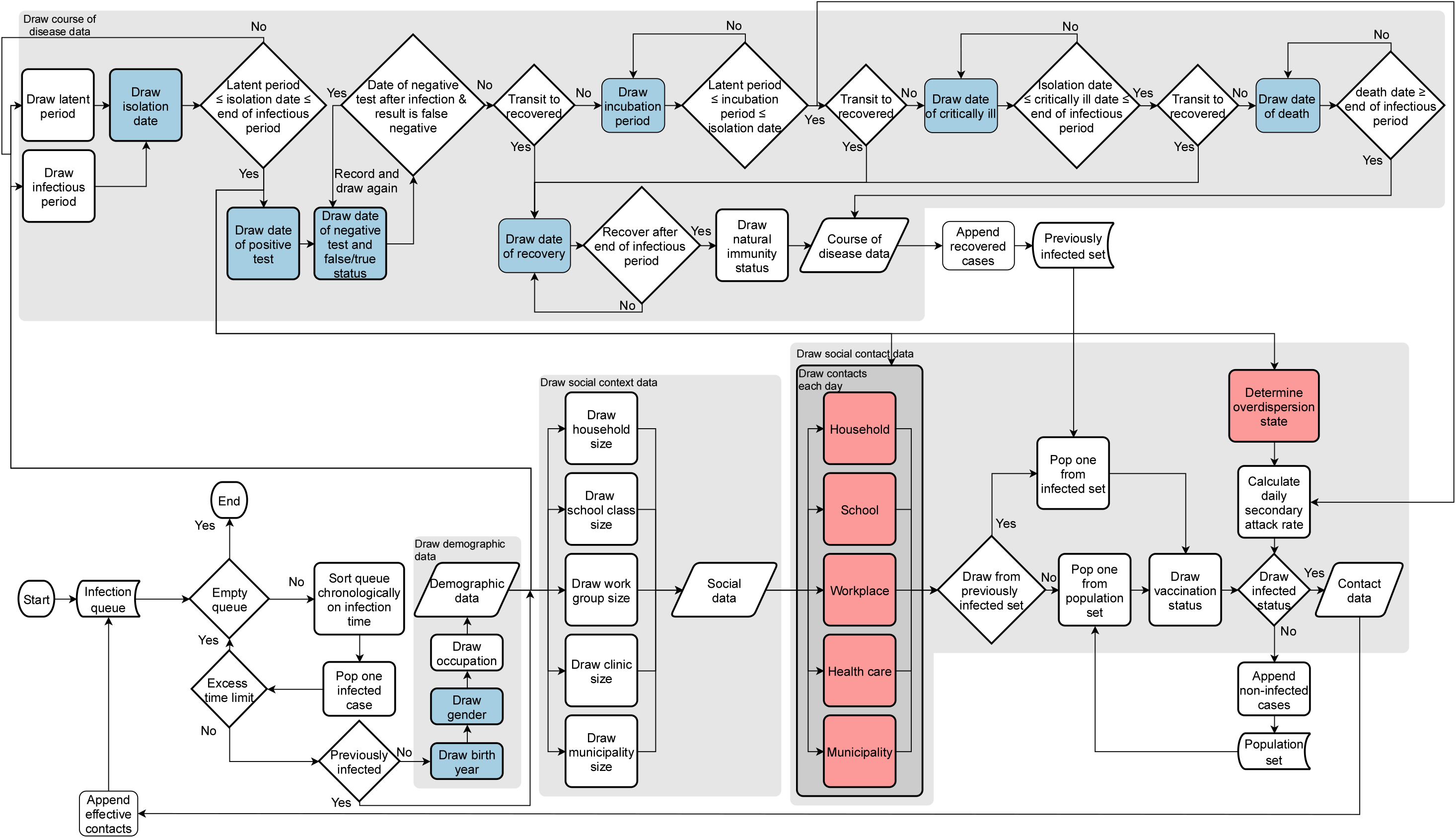
Flow chart of the data synthesis process. The process begins with an infection queue specifying the case IDs and infection time and ends when the queue is empty or the time limit has been reached. The final results are demographic data, social data, contact data, and course of disease data. Blue-colored boxes indicate that Taiwanese data contains parts of those data. Unknown parameters are indicated by red boxes. Reproduced from a manuscript under CC BY license [9].

## II. Method

Our data synthesis algorithm consists of four main modules: draw demographic data, draw social context data, draw course of disease data, and draw contact data. An outbreak is simulated by inputting an individual in the infection queue. Each individual in the infection queue is assigned a case ID, infection time, and previously infected status. As long as the queue is not empty and the outbreak time does not exceed the time limit, the process will keep synthesizing source cases’ detailed information.

### A. Draw demographic and social context data

Demographic data includes a person’s age, gender, and occupation. The social context data are categorized into five different layers, which are household, school, workplace, health care, and community. Based on the size of social context data randomly selected from Taiwanese social data, contacts will be drawn each day for each layer.

### B. Draw course of disease data

The latent period is defined as the time from the infection date to the beginning of the infectious period and the incubation period is defined as the time from infection to symptom onset. Each subject has different infectious periods (the grey shaded area in Figure 1), typically starting a few days before onset-of-symptom and ending when the viral load is low. The generation time is defined as the time between infection of the infector-infectee pair and the serial interval as the symptom-onset interval between the infector-infectee. If the case is confirmed and moved to monitored isolation before the end of the infectious period, then the infected person cannot infect other citizens, reducing the spread. Some cases may develop severe symptoms or even die, while most cases recover from the disease and become immune. Eventually, they lose their immunity and return to being susceptible.

The latent period, infectious period, and isolation time were simulated. We set the isolation date to be less than the sum of the latent period and infectious period. Since every case in our training data set has been tested and isolated, we have implemented this restriction, which can be relaxed later.

A subject can either move to state symptomatic or recovered after infection. If the person is symptomatic, the person can either move to the critically ill or recovered state. If the person is critically ill, the person can either recover or die. In each transition, we draw a time of progression and check if the time agrees with typical disease progression. Natural immunity was also drawn, and it affected the attack rate. The course of disease is saved in the infected matrix so that we can access all dates of disease progression for any infected person. Our algorithm for drawing course of disease data is shown in Algorithm 1. Since we don’t simulate the transition for each course of disease state each day but instead simulate the transition time directly, the computation cost is reduced.

### C. Draw contact data

The contact each day for each layer was synthesized by input social size, course of disease (incubation period and isolation period), and contact probability. The source case can only have valid contacts during infection to isolation and each day had a certain contact probability. Based on the assumption that human behaviour of close contacts would change a few days after symptom-onset, following an exponential trend, we defined the contact probability function *f*_*p L*_ (*t*) for the case of no contact in the previous day by combining two logistic functions.

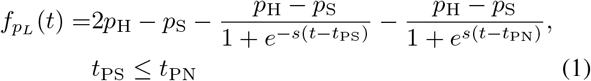

where *p*_H_ and *p*_S_ are the contact probability when healthy and symptomatic. *s* is the steepness. *t*_PS_ and *t*_PN_ are the phases relative to symptom-onset 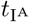 for symptom starting and toward recovering back to the normal contact pattern.

After the draw contacts each day process, the contact date for each contact in each layer is simulated. The source case might have multiple contacts either within the previously infected set or the population set. The vaccination status is drawn for each contact. All the contact types, vaccination status, natural immunity, and overdispersion state will affect the calculation of the daily secondary attack rate. With the secondary attack rate and contact dates, we can decide which date the contact gets infected, *i*.*e*. the effective contact date. The effective contact list was then sorted by infection time and appended back to the infection queue to start the next iteration. Our algorithm for drawing contact data is shown in Algorithm 2.

### D. Energy statistics

We apply energy distance statistics to quantify how different our synthetic dataset (Monte-Carlo events) is from the observed Taiwanese dataset. The null hypothesis is that both multidimensional data come from the same distribution. The idea is to make the synthetic dataset similar to the observed Taiwanese data.

Consider the two datasets 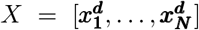 and 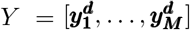. Each of the ***x***^***d***^ and ***y***^***d***^ is a *d* dimentional vector.

The energy distance is defined as

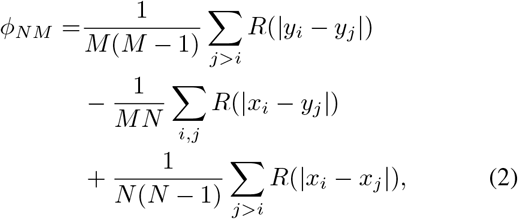

where *R*(·) is the distance function. We apply the logarithmic distance function

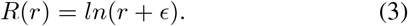

The small positive constant *ϵ* is set to be 10^−16^. Because our observed data is a sparse matrix, *i*.*e*. containing many NaN values, we remove the NaN and calculate the euclidean distances. The missing values are ignored in order to not distort the distribution.

The test data matrix of Taiwanese data contained features such as age, gender, time from infection to symptomatic, time from infection to recovery, time from symptomatic to critically ill, time from symptomatic to critically ill, time from critically ill to recovered, time from critically ill to death, time from the first negative test to confirmed, time from symptomatic to confirmed, and size of unique contacts. The asymptomatic date was not considered a test feature since it was an oversimplified estimated value and could cause the distortion of the real distribution.

### E. Optimization by Firefly algorithm

Since our model is complex with non-linear behaviour and has multiple local minima, gradient-based methods cannot explore the parameter space and produce good parameter estimates. Therefore, we implement the Firefly algorithm, which is a soft-computing optimization method, capable of producing a result closer to the global minimum without being trapped in the first local minimum.

As a cost function, we use the residual sum of square (RSS) of the synthesis secondary contact number and infection number each day versus the observation by Cheng *et al*. [10]. The contact number each day is grouped as *<* 0, 0 − 3, 4 − 5, 6 − 7, 8 − 9, and *>* 9 days from onset to the first exposure. Since Cheng *et al*. did not provide the specific layers’ statistics, we fit the sum of the contacts of our school, workplace, and municipality layers to the sum of Cheng *et al*.’s ‘nonhousehold family’ and ‘others’ layers. The energy distance (*E*) is also considered in the optimization to minimize the distance between Taiwan CDC data and our synthetic dataset.

#### Algorithm 1

Draw course of disease data

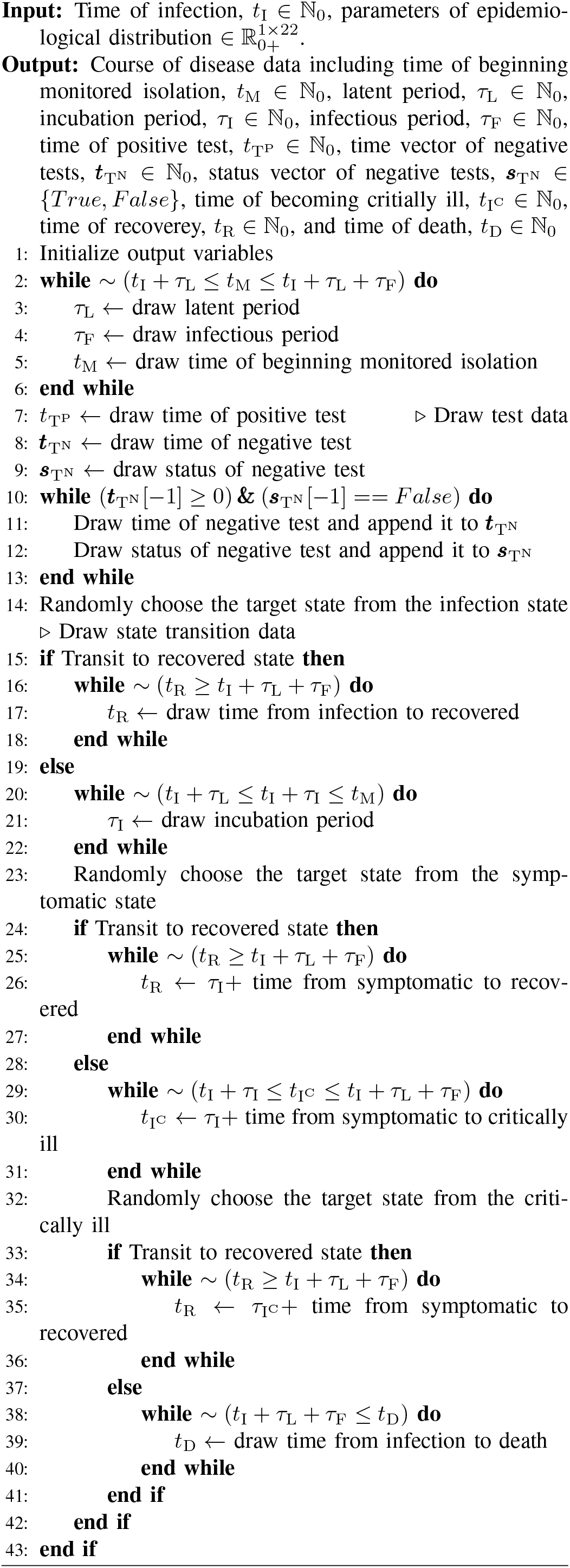

#### Algorithm 2

Draw contact data

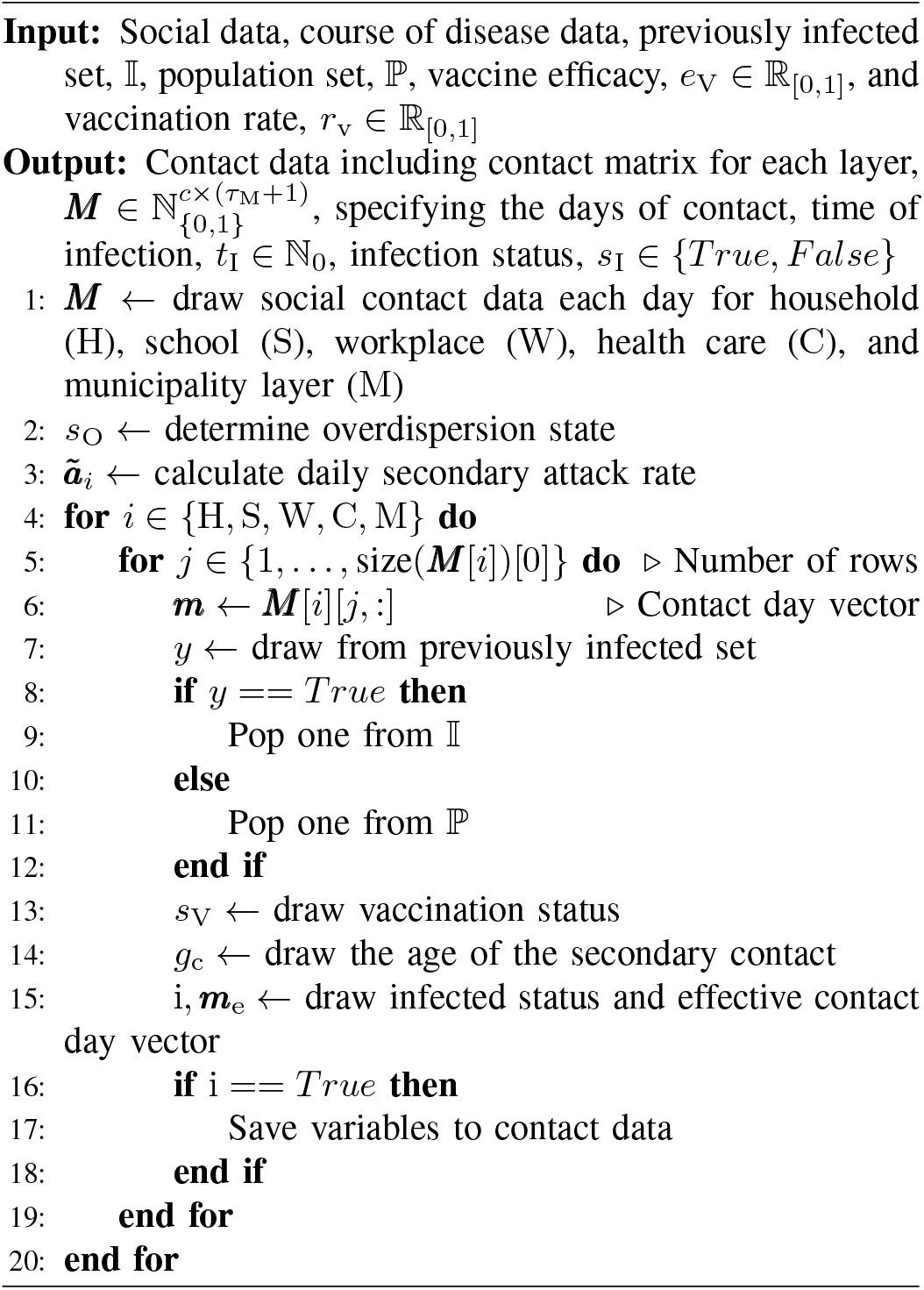

Finally, the cost function is defined as,

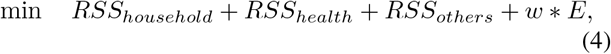

where *w* is a weight set to be 1000, since we observed that *E* was 3 order of magnitude smaller than *RSS*.

Optimization was performed on a Linux server with an AMD Ryzen Threadripper 1950x 16-core processor, 62G RAM, and Ubuntu 7.5.0, taking approximately 18 hours to optimize 194 parameters with a population size of 50 and 20 generations. Synthetic data and profile files were created on a Windows laptop with an Intel Core i7-7700HQ CPU, 16G RAM, and Windows 10. The Python Cprofiler and Snakeviz were used for iterative improvement of the algorithm, which included optimizing the numpy loading function, replacing the numpy random choice function with the Python random choice function, splitting if statements, and applying preallocation.

## III. Result

We report the difference between population and course of disease data, the optimization of the parameters, and generation of synthetic data.

### A. Measurement error of population data

We first compare population statistics to the seroprevalence study conducted by Roxhed *et al*. in Stockholm, Sweden. The study utilized tests on 878 randomly sampled individuals and found that the seroprevalence was 12.5% [11]. This indicated ∼ 150,000 inhabitants would have been infected with SARS-CoV-2 while the reported PCR-confirmed cases were just ∼ 13,000 during the study period. US, Taiwanese, and Iranian studies also showed that the estimated prevalence was much higher than the reported cases [12]–[14], see Figure 3.

**Fig. 3:**
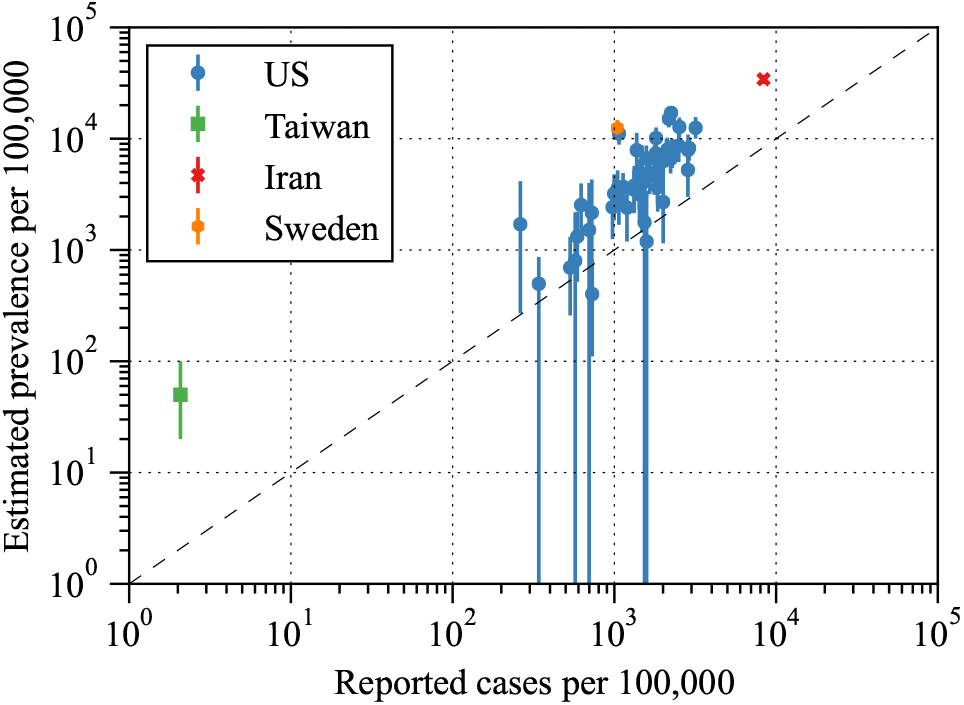
Seroprevalence estimates vs reported cases. Data is extracted from US, Taiwan, Iran, and Sweden [11]–[14].

The WHO estimate of excess deaths up until December 2021 was around 15 million compared to the reported COVID-19 deaths of 5.4 million. Furthermore, according to *The Economist*, the world estimated central excess death on Dec. 27, 2021, was 14.4 million with a lower bound of 12.9 million and an upper bound of 19 million, while the reported confirmed COVID-19 deaths was 5.5 million. The excess death was much higher than the reported death indicating that the death measurement was also problematic [15], [16].

We collected the estimated reproduction number of population-based models (Figure 7). The estimats of *R*_0_ were widespread from 0.17 to 4.5 with confidence interval ranging from 0 to 12.26.

### B. Comparison of synthetic population data and individual data

We synthesized 10 subjects’ course of disease data and highlight the increased informativeness of individual data compared to population data. Figure 5A displays the daily cumulative number of different states, such as symptomatic, confirmed, critically ill, recovered, and dead, which are commonly obtained from open databases. The cumulative number of infected individuals is difficult to measure and not present in any open database. Figure 5B presents the branching process of the ten cases, similar to Figure 1. Effective contacts between cases are marked as diamonds and only occur during the infectious period. Note that some cases start the infectious period before symptom onsets such as cases I2, I5, I6, and I10. The detailed contact information of each subject is plotted in the daily contact network (Figure 5C) and the contact network in various social contexts (Figure 5D). Uninfected contacts are labelled by integers and represented as small nodes, while infected cases are labelled by I+integer, where ‘I’ stands for ‘infected’. Figure 5C shows that uninfected subjects can have multiple days of contact with the source case without becoming infected. For example, subject 0 has contact with subject I1 on days 14 and 15. The effective contact is shown in the example of I2 being infected by I1 on day 19. Due to the complexity of the network, we only show the contacts until index 10 and the timelimit 23 days. Figure 5D presents the contact network visualized using Cytoscape version 3.9.1, with different layers coloured differently. In this example, there is no contact in the school layer. Different uninfected subjects can have contact with multiple infected cases, such as subject 83 having contact with I5 and I6.

**Fig. 4:**
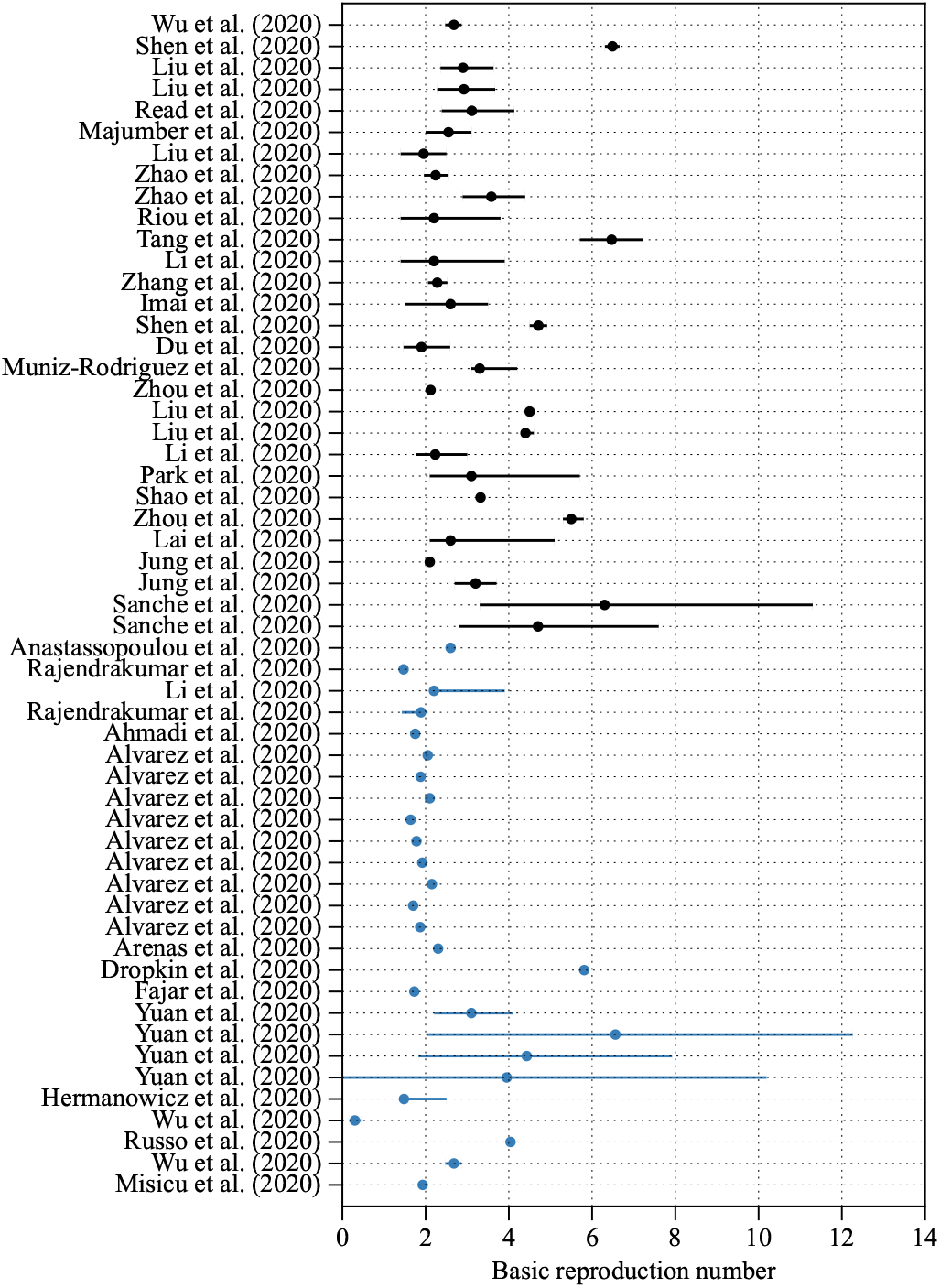
Estimates of *R*_0_ using population-based models. The black dots were extracted from [17] and the blue dots were extracted from [18]–[31].

**Fig. 5:**
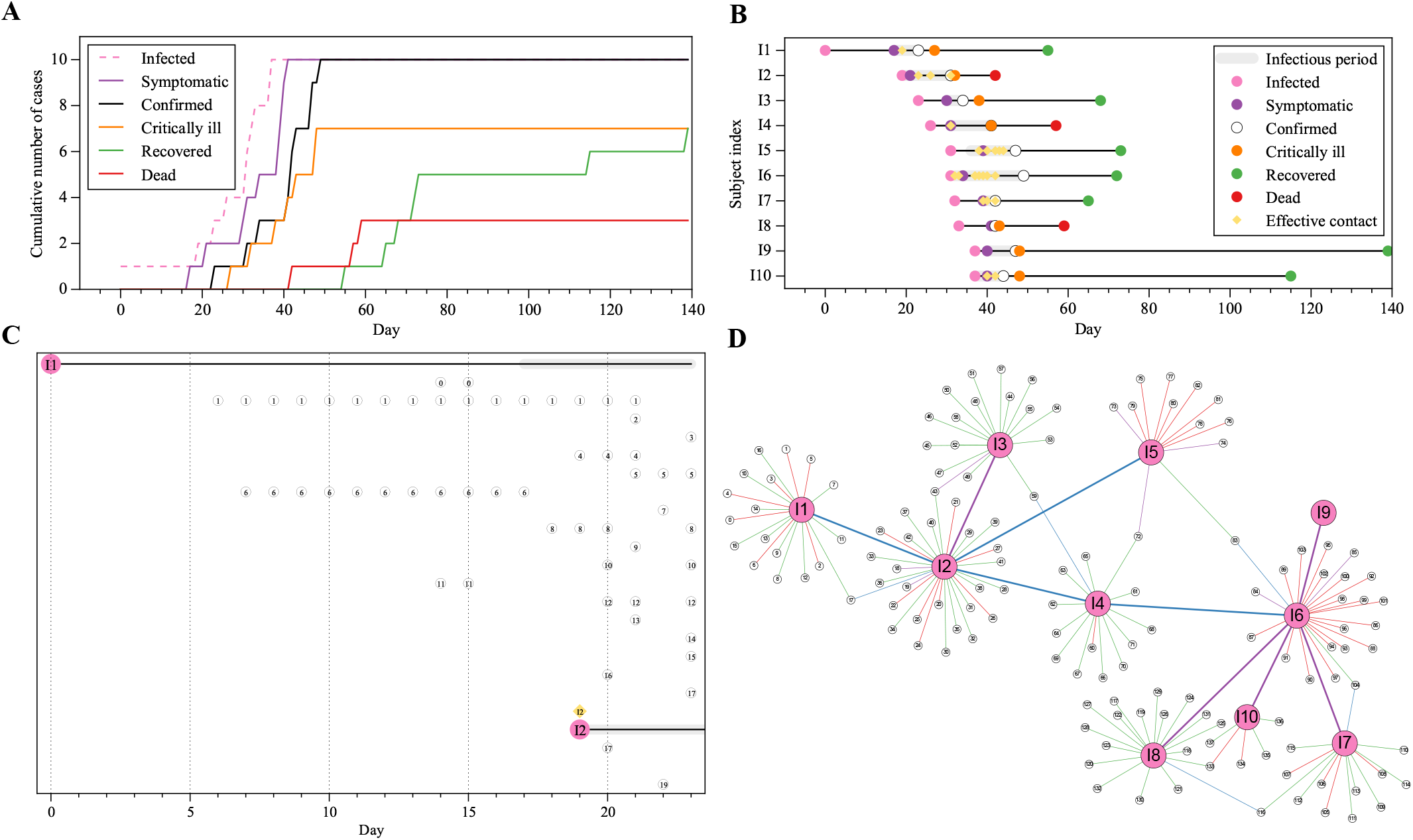
Comparison of population data and individual data. Figure A presents the daily cumulative count of symptomatic, confirmed, critically ill, recovered, and dead, which are obtainable from open databases. The daily number of infected individuals is indicated by a dashed line, reflecting the difficulty to measure it. Figure B depicts the course of disease for the ten synthesized cases, including the specific dates of state transitions and the infectious period, during which the subject is capable of infecting others. Figure C shows the daily count of uninfected contacts of source cases I1 and I2, labelled as integers, each representing a different day of contact with the source case in the same row. Figure D displays the contact network of the infected cases (large nodes) and uninfected contacts (small nodes), with different types of contacts distinguished by different colours. Household contacts are blue, workplace contacts are green, healthcare contacts are red, and other contacts are purple.

Based on Taiwanese seroprevalence research [13], we compare population data with median detection rates of 0.04 and lower and upper bounds of 0.02 and 0.1. The death detection rate was set to be 0.4 based on *The Economist*’s estimation [15]. The results of our Monte-Carlo simulation with 1,000 iterations are displayed in log scale in Figure 6. The actual cumulative number of confirmed cases on day 150 was 1454, while the median detected number of confirmed cases on the final day were 154, 68, and 39, respectively for 0.1, 0.04, and 0.02.

**Fig. 6:**
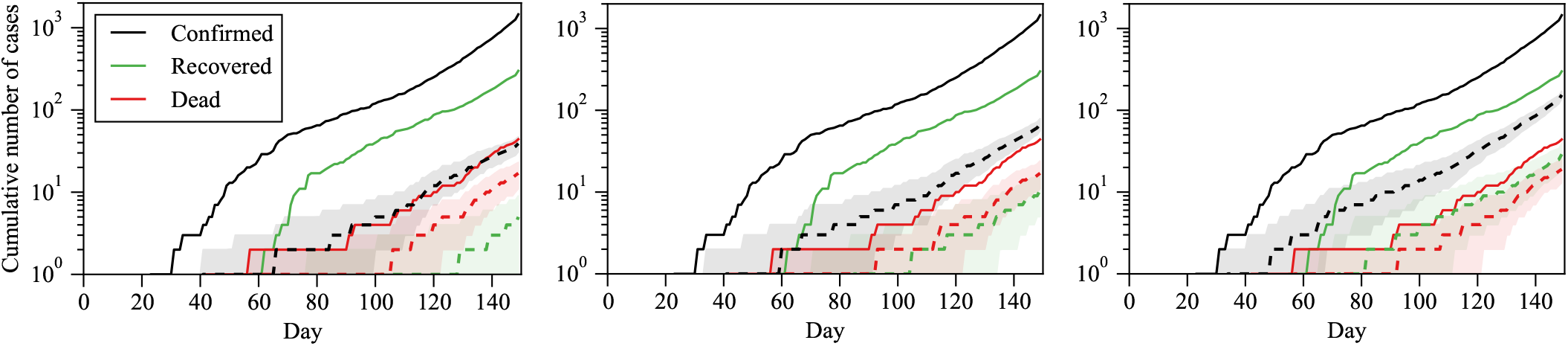
Illustration of the impact of failure to detect infected individuals on population statistics. The true cumulative number of cases (solid line) and the 95% confidence interval (shaded area) with median (dashed line) are based on 1,000 Monte Carlo simulations assuming detection rates of 0.02 (left), 0.04 (middle) and 0.1 (right) for confirmed and recovered cases and 0.4 for deaths.

*R*_0_ values for population data were estimated using the least-squares method under the SIRD model structure [18]. This was a widely adopted method during the beginning of the COVID-19 pandemic, with over a thousand citations. Note that the SIRD approach did not take into account factors such as the incubation period, heterogeneous contact transmission, age, and so on, which could cause bias when estimating *R*_0_. Furthermore, this method is not reliable after the early stages of an epidemic.

We also estimated *R*_0_ values directly from the contact network by averaging the effective contact number for each subject. We reported the mean and median values of the *R*_0_ estimates in Table I and Figure 7. Since *R*_0_ is an average number and the mean values tend to be higher than the median values, which can lead to faster reaction times in practice, we estimated the baseline *R*_0_ by averaging the effective contact number over a total of 3,718 simulated source objects. Additionally, we observed a trend where the uncertainty range tended to decrease as the number of subjects increased. For instance, our findings indicate that the uncertainty range for 100 subjects was comparable in magnitude to the estimates obtained from population data.

**TABLE I:**
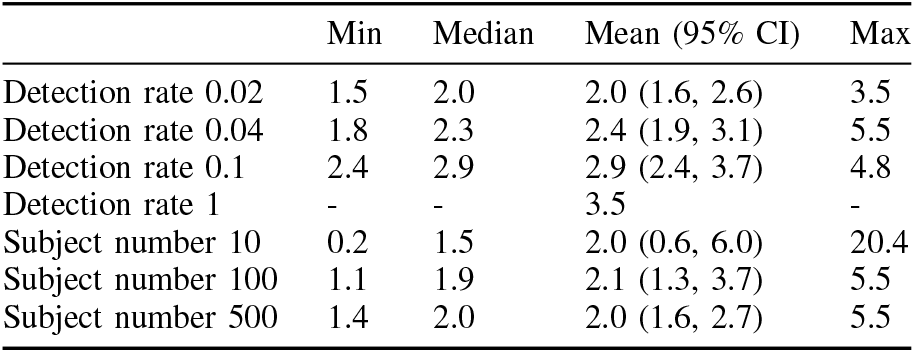
*R*_0_ estimates based on population data (first four) and contact networks (last three). The estimate of all subject based on the average of the effective contact numbers of 3,718 subjects is 2.1.

### C. Synthesization and validation of individual data

In Figure 8 each firefly is initially scattered across the cost landscape marked as red dots, representing a diverse range of solutions. As the algorithm progresses, the fireflies undergo a process of exploration and they move towards the regions of higher cost and then move toward optimal results.

**Fig. 7:**
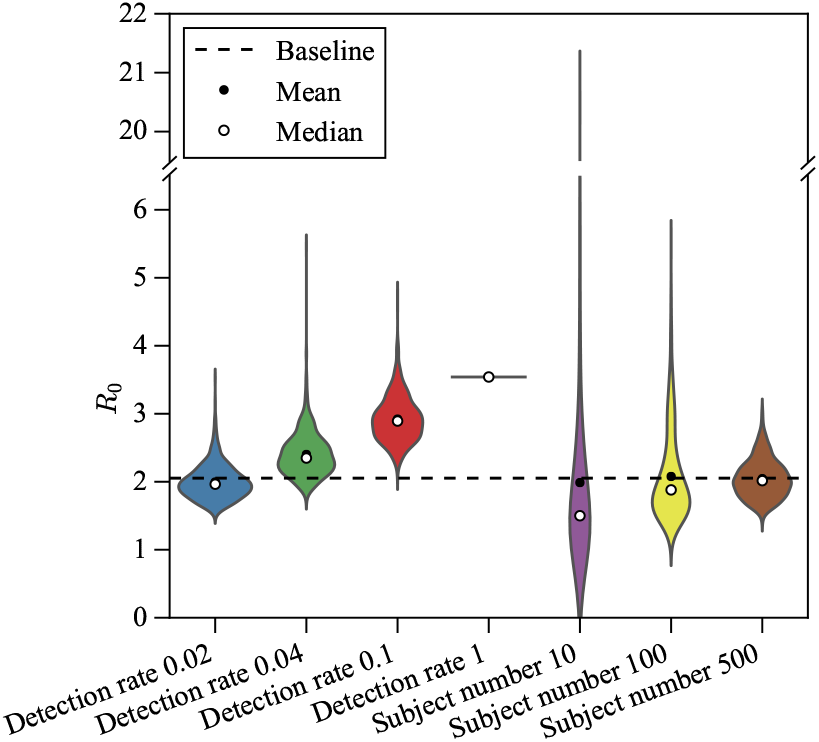
Violin plot of *R*_0_ values estimated by SIRD least-square method (left four) and averaging effective contacts (right three). The Baseline was calculated by averaging effective contacts of 3,718 source subjects.

**Fig. 8:**
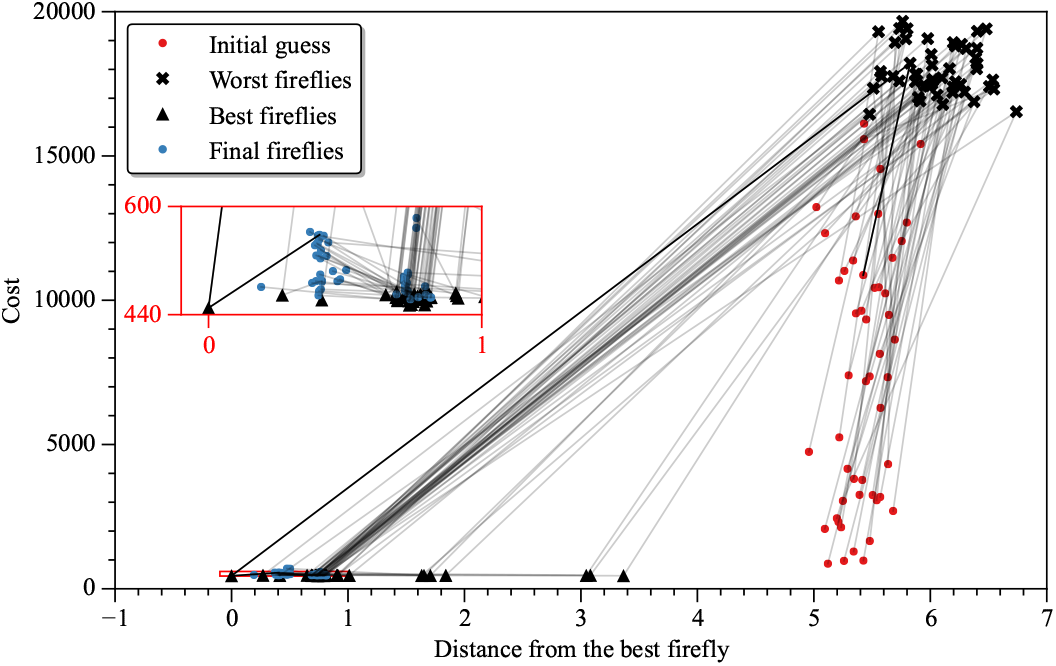
Firefly distance. The plot indicates that the system has multiple local minima. We saved each firefly’s initial guess, worst cost, best cost, and final cost. The red rectangle shows the zoomed-in plot around the final fireflies.

We compared our synthetic dataset with the Taiwanese data shown in Figure 9. The simulation result stays in the 95% confidence interval of the Taiwanese data except for around day 24 and day 34.

**Fig. 9:**
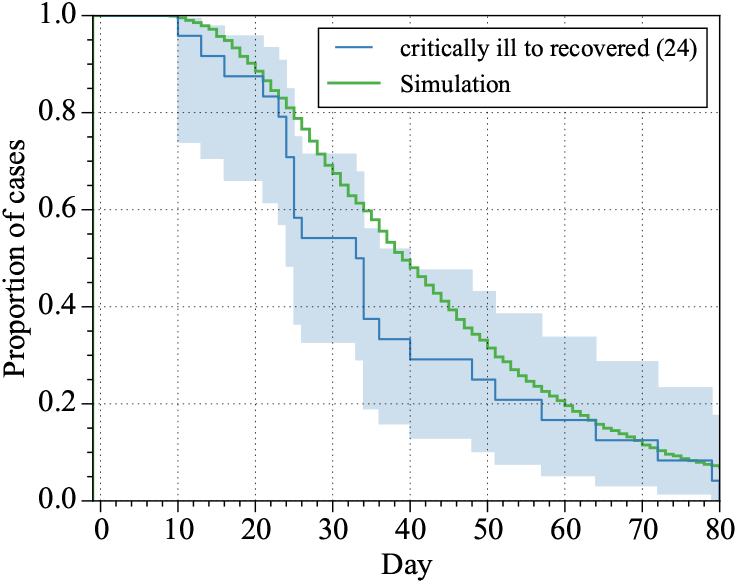
Proportion of cases remaining critically ill each day after transition. The blue lines are extracted from the Taiwanese data and the green line is based on 10,000 simulations.

Furthermore, the energy distance between our synthetic dataset and the Taiwanese data was 0.19 with p-value 0.67 meaning that the null hypothesis that both datasets are from the same distribution cannot be rejected.

### D. Algorithm optimization

The running time of the core code was iteratively improved from 0.011s to 0.007s. The detailed profile files are shown in Figure 10.

**Fig. 10:**
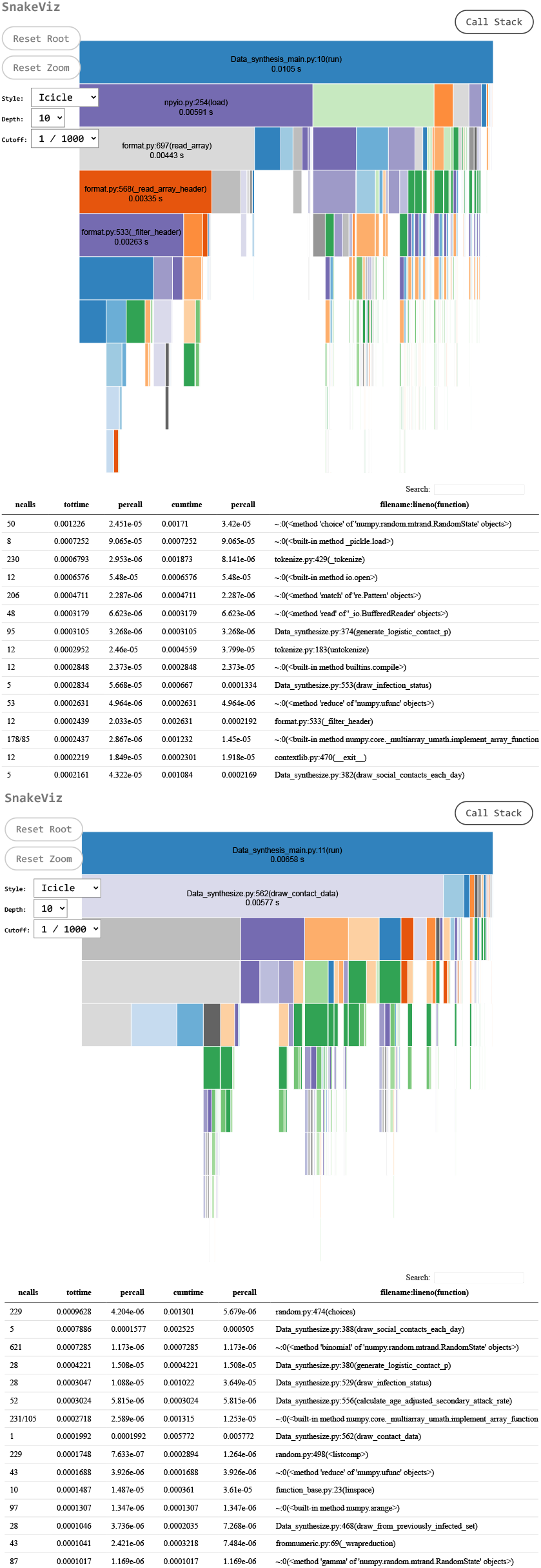
Comparison of the core code Python profile file of `Data|_synthesis_main .py`, before optimization (top) and after (bottom).

## IV. Discussion and conclusion

In section III-A, we demonstrate the unreliability of commonly used measurements such as the number of infections, deaths, and *R*_0_. Despite this, all state-of-the-art COVID-19 models rely on population data, without utilizing individual-level information about the course of disease and contact networks, even in agent-based models.

In this article, we take a pure empirical data based approach to epidemiology. Only when data is not available do we use relationships derived from past studies, such as statistical distribution or parameter values. The open Taiwanese COVID-19 data has limited individual information, making analysis and modeling challenging. To overcome this, we synthesized additional individual data based on the collected data and COVID-19 epidemiological parameters. We synthesized ten cases and compared them with population data in Figure 5. Daily cumulative case is an oversimplification by ignoring the information about each subject’s different course of disease, daily contact network, and contact type. We also demonstrate the impact of the detection rate on population measurement in Figure 6.

To validate the effectiveness of the firefly optimization in balancing exploration and exploitation, we plot the cost versus distance from the best firefly to the others in Figure 8. All fireflies converged towards the steady state but have not yet reached it. This indicates that the number of generation need to be increased. Improving the core code’s efficiency is critical, as it is repeatedly executed hundreds of thousands of times during the optimization process. Our simulation result shown in Figure 9 stays in the 95% confidence interval for most of the states and days except for the critically ill state. This also suggests that our parameters have not been optimized to the global minimum. Nonetheless, our model is still useful for our analysis.

In the epidemiological modelling field, we are the first to develop a synthetic dataset for disease model development. These ideas have been validated in the systems biology field, *e*.*g*. synthetic gene expression datasets for inference of gene regulatory networks and published in top journals [32], [33]. The algorithm we developed for generating synthetic COVID-19 datasets enables benchmarking of epidemiological models. After better optimization, our model is expected to give more accurate forecasts to inform decision-makers and save lives in future pandemics.

## Data Availability

All data produced in the present study are available upon reasonable request to the authors

## Appendix

### A. Taiwan COVID-19 individual data collection and preprocessing

The Taiwanese COVID-19 individual data of confirmed cases and the daily summary data were collected from the following open source databases: Taiwan CDC Open Data Portal, Regents of the National Center for High-performance Computing (COVID-19 Dashboard), United Daily News (Visualization of contacts of Taiwan COVID-19 cases), Taiwan Centers for Disease Control (CDC) press release, and Taiwan Centers for Disease Control press conference. We use the quantitative structured data that we composed based on public sources in Taiwan. We fit Gamma distributions to all state transition days. Statistics from other studies were used for parameters with insufficient data, such as the latent period.

### B. Data preprocessing

We collected individual course of disease data from 2020-02-05 to 2020-11-09 covering the first outbreak in Taiwan. The second outbreak started from 2021-01-12 to 2021-02-09 and the third outbreak started on 2021-04-20. Both the second and third outbreaks contain the alpha variant, which was estimated to have a 1.44-fold higher infection probability and 57% higher reproduction number [34]. Since the alpha variant has a different attack rate, and the open-source Taiwanese data does not specify alpha cases, we did not use the data for modelling. Moreover, Taiwan CDC stopped providing individual case data starting on 2021-05-15 due to the sudden daily increase from 29 cases to 185 cases, which makes it challenging to collect detailed data.

The confirmed case data contain 579 samples with 64 features including travel history, age, gender, nationality, the onset of symptom, confirmed date, symptoms, way of discovery, and contact types between cases. We categorize the contact type into groups, such as couples, parents, grandparents, siblings, family, friends, live together, flight, flight (nearby seat), travelling, school, car, coworker, hospital, hotel, Panshi combat ship, Coral Princess, and others. Some cases also contain ICU date, recovery date, and death date, depending on the availability of sources. The daily summary data released by Taiwan CDC from 2020-01-21 to 2022-05-23 provided population data, including the number of suspected cases, excluded cases, abroad positive cases, local positive cases, Panshi ship positive cases, positive cases with unknown sources, deaths, recovery cases, and hospital quarantine cases. Contact information is not available for all the cases. But when available, Taiwan CDC reported the number of close contacts, contact dates, and case indexes of the contact.

## References

[1] N. Ferguson, D. Laydon, G. Nedjati Gilani, N. Imai, K. Ainslie, M. Baguelin, S. Bhatia, A. Boonyasiri, Z. Cucunuba Perez, G. Cuomo-Dannenburg et al., “Report 9: Impact of non-pharmaceutical interventions (npis) to reduce covid19 mortality and healthcare demand, “ Imperial College COVID-19 Response Team, 2020.

[2] J. Hellewell, S. Abbott, A. Gimma, N. I. Bosse, C. I. Jarvis, T. W. Russell, J. D. Munday, A. J. Kucharski, W. J. Edmunds, F. Sun et al., “Feasibility of controlling covid-19 outbreaks by isolation of cases and contacts, “ The Lancet Global Health, vol. 8, no. 4, pp. e488–e496, 2020.

[3] IHME, “Modeling covid-19 scenarios for the united states, “ Nature medicine, vol. 27, no. 1, pp. 94–105, 2021.

[4] A. Srivastava, T. Xu, and V. K. Prasanna, “Fast and accurate forecasting of covid-19 deaths using the sikjα model, “ arXiv preprint arXiv:2007.05180, 2020.

[5] A. Rodriguez, A. Tabassum, J. Cui, J. Xie, J. Ho, P. Agarwal, B. Adhikari, and B. A. Prakash, “Deepcovid: An operational deep learning-driven framework for explainable real-time covid-19 forecasting, “ in Proceedings of the AAAI Conference on Artificial Intelligence, vol. 35, no. 17, 2021, pp. 15393–15 400.

[6] S. Zheng, Z. Gao, W. Cao, J. Bian, and T.-Y. Liu, “Hierst: A unified hierarchical spatial-temporal framework for covid-19 trend forecasting, “ in Proceedings of the 30th ACM International Conference on Information & Knowledge Management, 2021, pp. 4383–4392.

[7] D. Wu, L. Gao, X. Xiong, M. Chinazzi, A. Vespignani, Y.-A. Ma, and R. Yu, “Deepgleam: a hybrid mechanistic and deep learning model for covid-19 forecasting, “ arXiv preprint arXiv:2102.06684, 2021.

[8] C. C. Kerr, R. M. Stuart, D. Mistry, R. G. Abeysuriya, K. Rosenfeld, G. R. Hart, R. C. Núñez, J. A. Cohen, P. Selvaraj, B. Hagedorn et al., “Covasim: an agent-based model of covid-19 dynamics and interventions, “ PLOS Computational Biology, vol. 17, no. 7, p. e1009149, 2021.

[9] Y.-H. Wu and T. E. Nordling, “An agent-based model for synthesizing COVID-19 course of disease data, “ Manuscript, 2023.

[10] H.-Y. Cheng, S.-W. Jian, D.-P. Liu, T.-C. Ng, W.-T. Huang, and H.-H. Lin, “Contact Tracing Assessment of COVID-19 Transmission Dynamics in Taiwan and Risk at Different Exposure Periods Before and After Symptom Onset, “ JAMA Internal Medicine, vol. 180, no. 9, p. 1156, sep 2020. [Online]. Available: https://jamanetwork.com/journals/jamainternalmedicine/fullarticle/2765641

[11] N. Roxhed, A. Bendes, M. Dale, C. Mattsson, L. Hanke, T. Dodig-Crnković, M. Christian, B. Meineke, S. Elsässer, J. Andréll et al., “Multi-analyte serology in home-sampled blood enables an unbiased assessment of the immune response against sars-cov-2, “ Nature Communications, vol. 12, no. 1, pp. 1–9, 2021.

[12] K. L. Bajema, R. E. Wiegand, K. Cuffe, S. V. Patel, R. Iachan, T. Lim, A. Lee, D. Moyse, F. P. Havers, L. Harding et al., “Estimated sars-cov-2 seroprevalence in the us as of september 2020, “ JAMA internal medicine, vol. 181, no. 4, pp. 450–460, 2021.

[13] H.-L. Ho, F.-Y. Wang, H.-R. Lee, Y.-L. Huang, C.-L. Lai, W.-C. Jen, S.-L. Hsieh, and T.-Y. Chou, “Seroprevalence of covid-19 in taiwan revealed by testing anti-sars-cov-2 serological antibodies on 14,765 hospital patients, “ The Lancet Regional Health-Western Pacific, vol. 3, p. 100041, 2020.

[14] M. Zamani, H. Poustchi, Z. Mohammadi, S. Dalvand, M. Sharafkhah, S. A. Motevalian, S. Eslami, A. Emami, M. H. Somi, J. Yazdani-Charati et al., “Seroprevalence of sars-cov-2 antibody among urban iranian population: findings from the second large population-based cross-sectional study, “ BMC public health, vol. 22, no. 1, pp. 1–10, 2022.

[15] S.-U. Solstad, “The pandemic ‘s true death toll, “ The Economist, 2021.

[16] V. Knutson, S. Aleshin-Guendel, A. Karlinsky, W. Msemburi, and J. Wakefield, “Estimating global and country-specific excess mortality during the covid-19 pandemic, “ arXiv preprint arXiv:2205.09081, 2022.

[17] Y. Alimohamadi, M. Taghdir, and M. Sepandi, “Estimate of the basic reproduction number for covid-19: a systematic review and meta-analysis, “ Journal of Preventive Medicine and Public Health, vol. 53, no. 3, p. 151, 2020.

[18] C. Anastassopoulou, L. Russo, A. Tsakris, and C. Siettos, “Data-based analysis, modelling and forecasting of the covid-19 outbreak, “ PloS one, vol. 15, no. 3, p. e0230405, 2020.

[19] A. L. Rajendrakumar, A. T. N. Nair, C. Nangia, P. K. Chourasia, M. K. Chourasia, M. G. Syed, A. S. Nair, A. B. Nair, and M. S. F. Koya, “Epidemic landscape and forecasting of sars-cov-2 in india, “ Journal of Epidemiology and Global Health, vol. 11, no. 1, p. 55, 2021.

[20] Q. Li, X. Guan, P. Wu, X. Wang, L. Zhou, Y. Tong, R. Ren, K. S. Leung, E. H. Lau, J. Y. Wong et al., “Early transmission dynamics in wuhan, china, of novel coronavirus–infected pneumonia, “ New England journal of medicine, 2020.

[21] A. Ahmadi, Y. Fadaei, M. Shirani, and F. Rahmani, “Modeling and forecasting trend of covid-19 epidemic in iran until may 13, 2020, “ Medical Journal of the Islamic Republic of Iran, vol. 34, p. 27, 2020.

[22] L. Alvarez, “A model to forecast the evolution of the number of covid-19 symptomatic patiens after drastic isolation measures, “ 2020.

[23] A. Arenas, W. Cota, J. Gómez-Gardenes, S. Gómez, C. Granell, J. T. Matamalas, D. Soriano, and B. Steinegger, “A mathematical model for the spatiotemporal epidemic spreading of covid19, “ MedRxiv, pp. 2020–03, 2020.

[24] G. Dropkin, “Covid-19 uk lockdown forecasts and r 0, “ Frontiers in public health, vol. 8, p. 256, 2020.

[25] M. Fajar and U. Padjadjaran, “Estimation of covid-19 reproductive number case of indonesia, “ Badan Pusat Statistik Indonesia, 2020.

[26] J. Yuan, M. Li, G. Lv, and Z. K. Lu, “Monitoring transmissibility and mortality of covid-19 in europe, “ International Journal of Infectious Diseases, vol. 95, pp. 311–315, 2020.

[27] S. W. Hermanowicz, “Forecasting the wuhan coronavirus (2019-ncov) epidemics using a simple (simplistic) model, “ MedRxiv, pp. 2020–02, 2020.

[28] P. Wu, X. Hao, E. H. Lau, J. Y. Wong, K. S. Leung, J. T. Wu, B. J. Cowling, and G. M. Leung, “Real-time tentative assessment of the epidemiological characteristics of novel coronavirus infections in wuhan, china, as at 22 january 2020, “ Eurosurveillance, vol. 25, no. 3, p. 2000044, 2020.

[29] J. T. Wu, K. Leung, and G. M. Leung, “Nowcasting and forecasting the potential domestic and international spread of the 2019-ncov outbreak originating in wuhan, china: a modelling study, “ The Lancet, vol. 395, no. 10225, pp. 689–697, 2020.

[30] L. Russo, C. Anastassopoulou, A. Tsakris, G. Bifulco, E. Campana, G. Toraldo, and C. Siettos, “Tracing day-zero and forecasting the fade out of the covid-19 outbreak in lombardy, “ Italy: a compartmental modelling and numerical optimization approach, 2020.

[31] S. Misicu et al., “Beginning of the end of the pandemic? a comparison between italy and romania. “ medRxiv, 2020.

[32] I. Cantone, L. Marucci, F. Iorio, M. A. Ricci, V. Belcastro, M. Bansal, S. Santini, M. Di Bernardo, D. Di Bernardo, and M. P. Cosma, “A yeast synthetic network for in vivo assessment of reverse-engineering and modeling approaches, “ Cell, vol. 137, no. 1, pp. 172–181, 2009.

[33] D. Marbach, J. C. Costello, R. Küffner, N. M. Vega, R. J. Prill, D. M. Camacho, K. R. Allison, M. Kellis, J. J. Collins, and G. Stolovitzky, “Wisdom of crowds for robust gene network inference, “ Nature methods, vol. 9, no. 8, pp. 796–804, 2012.

[34] C.-Y. Hsu, J.-T. Wang, K.-C. Huang, A. C.-H. Fan, Y.-P. Yeh, and S. L.-S. Chen, “Household transmission but without the community-acquired outbreak of COVID-19 in Taiwan. “ Journal of the Formosan Medical Association = Taiwan yi zhi, may 2021.

